# The Burden and Genomic Characterization of *Shigella*-Associated Diarrhea in Children Under Five in Lusaka, Zambia: A Prospective Cohort Study

**DOI:** 10.64898/2026.05.14.26353268

**Authors:** Mwelwa Chibuye, Vanessa C Harris, Jaime Brizuela, Samuel Bosomprah, Michelo Simuyandi, Kapambwe Mwape, Suwilanji Silwamba, Fraser Liswaniso, Kennedy Chibesa, Sam Miti, Gonçalo Piedade, Charlie C Luchen, Caroline C Chisenga, Daniel R Mende, Constance Schultsz, Roma Chilengi

## Abstract

**Background:** *Shigella* is a leading cause of childhood diarrhea in low- and middle-income countries and is increasingly resistant to first-line antibiotics. We conducted a surveillance study to determine the incidence, genomic characteristics, and AMR profiles of *Shigella* infections in children under five with moderate to severe diarrhea (MSD) in Lusaka, Zambia.

**Methods:** Between 15 September 2020 and 30 November 2021, a prospective cohort study of 1,400 children under five was enrolled during a community census in a peri-urban setting and passively followed for 9.5 months for MSD. During enrollment, socio-demographic data were collected using electronic questionnaires, while clinical data were collected through the DHIS platform. The main outcome, *Shigella* in diarrheal stool in under 5 children, was detected using culture and Loop-mediated Isothermal Amplification (LAMP) targeting the ipaH gene. Cox proportional hazards models were used to assess the incidence and risk factors of *Shigella* (ipaH) infections. Whole-genome sequencing (WGS) was used to characterize the genomic diversity and antimicrobial resistance genes, complemented by phenotypic antibiotic susceptibility testing.

**Results:** There were 230 first episodes of *Shigella* over a follow-up time of 9,581.7 child-months, yielding an incidence of 24.0 (95% CI 21.1-27.3) cases per 1,000 child-months, with the highest incidence among 2 to 3-year-olds. The key risk factors identified were the water source (p=0.025) and age group (p=0.014). Genotypic characterization revealed 10 *S. flexneri*, 9 *S. sonnei*, and 3 *S. boydii*. The *S. sonnei* isolates formed two clusters, differing in virulence factors and plasmid profiles, indicating two possible circulating strains. *Shigella* isolates exhibited phenotypic and genotypic multidrug resistance, including against trimethoprim, aminoglycosides, and beta-lactams. Plasmid-mediated quinolone resistance (qnrS1) was identified in four *S. flexneri* isolates, with these genes located on the IncFIB(K) plasmid, highlighting the potential for horizontal transmission and spread of quinolone resistance in this region. No phenotypic and genotypic resistance to macrolides, the first-line treatment for Shigella in Zambia, was observed.

**Interpretation:** We report a high burden of *Shigella* with multidrug resistance, including resistance to fluoroquinolones. These findings highlight the increasing resistance of *Shigella* to first-line antibiotics and underscore the importance of developing safe and effective vaccines, improving WASH conditions, and ongoing AMR surveillance.

**Funding:** The EDCTP2 program, supported by the European Union, the Faculty for the Future Foundation (FFTF), the Netherlands Organization for Health Research and Development (ZonMw), and Health-Holland AMR-Global, Gloria, and Track-AMR.

**Research in context:** *Evidence before this study:* Despite *Shigella* being a leading cause of bacterial diarrheal mortality globally, there is a critical lack of up-to-date data on its burden and associated antimicrobial resistance (AMR) in high-risk settings, such as sub-Saharan Africa (SSA). We searched PubMed for studies published between 2000 and May 2025, using the terms “*Shigella*,” “antimicrobial resistance,” and “sub-Saharan Africa.” Only three studies (GEMS, MAL-ED, VIDA) involving extensive surveillance in SSA countries were identified, all conducted before 2018. None integrated disease burden, genomic characterization of circulating strains, and both phenotypic and genotypic AMR profiling. In Zambia, we found no published surveillance data on *Shigella* or resistance patterns in children.

*The added value of the study:* We present an up-to-date, integrated assessment of the burden, genomic diversity, and AMR profiles among *Shigella* isolates in children under five in SSA, specifically in a high-risk peri-urban setting in Lusaka, Zambia. By prospectively following a large cohort of 1,400 children and combining culture with WGS, we provide detailed insights into the disease burden and epidemiology of circulating *Shigella* strains and the relevance of candidate vaccine antigens. We reveal a high prevalence of multidrug resistance (MDR), including plasmid-mediated and phenotypic resistance to ciprofloxacin, the first-line treatment for *Shigella*. We further complement genotypic AMR with phenotypic AMR testing to predict potential resistance genes while measuring antibiotic susceptibility in real clinical settings.

*Implications of all the available evidence:* We demonstrate that *Shigella* is genomically diverse and an important etiology of moderate to severe gastroenteritis in Zambian children. Risk factors include increasing age and poor WASH. We observed plasmid-mediated resistance to ciprofloxacin and MDR, which threatens the efficacy of current *Shigella* treatment and risks population-level AMR spread. These results highlight the need for improved WASH, antibiotic stewardship, and the development of effective vaccines, supported by ongoing genomic surveillance, to facilitate disease monitoring, inform treatment guidelines, guide vaccine antigen selection, and inform evidence-based antibiotic stewardship.

## Introduction

*Shigella* is one of the leading causes of diarrheal disease and the second leading cause of diarrheal mortality in children under five, resulting in an estimated 81,800 diarrheal deaths and 7·34 million diarrheal disability-adjusted life-years (DALYs), with the highest mortality reported in low- and middle-income countries (LMICs). ^1^ It has a low infectious dose of 10 to 100 organisms and is endemic in resource-limited settings where frequent fecal exposures occur due to inadequate water, sanitation, and hygiene (WASH) and deficient healthcare infrastructure.^2^ Despite this, data from high-burden regions, including sub-Saharan Africa (SSA), is limited. Only three studies ^3–5^ have conducted comprehensive surveillance involving SSA to describe the burden of *Shigella*, all conducted before 2018, leaving a major public health knowledge gap. Up-to-date and strengthened *Shigella* surveillance could help justify investments in disease prevention and control, and is crucial for understanding disease burden, detecting outbreaks, and early identification of emerging antimicrobial resistance (AMR).

AMR limits treatment options for *Shigella*, increases morbidity and mortality, and increases healthcare costs.^6^ There has been a marked rise in *Shigella* infections with antimicrobial resistance (AMR) to first-line antibiotic therapies, including fluoroquinolones (mainly driven by mutations in the gyrase gene) and macrolides, especially in Asia, where resistance can vary from 3-70% for ciprofloxacin and 10-59%, for azithromycin.^7^ The rise in fluoroquinolone and macrolide resistance in Asia highlights the need to monitor changes in AMR in sub-Saharan regions, where AMR surveillance is limited and antibiotic usage differs markedly by country.

Although *Shigellosis* is a notifiable disease in Zambia and other SSA countries, testing for *Shigella* in diarrheal stool is not routine, and antibiotic treatment is mostly empiric based on the presence of blood in the stool. Vaccines against *Shigella* could be a cost-effective intervention by protecting children from infections, reducing antibiotic use, and mitigating the spread of disease.^8^ However, licensed *Shigella* vaccines are currently unavailable, though several candidate vaccines are under development. Reliable background data on the incidence and prevalence of AMR could guide antibiotic stewardship and vaccine rollout across SSA, yet up-to-date burden estimates are lacking. This paper describes moderate-to-severe diarrhea (MSD) with *Shigella* etiology in children under five in Lusaka, Zambia. We use whole-genome sequencing (WGS) to characterize circulating *Shigella* strains and determine their antimicrobial resistance profiles. This study provides important baseline data for future investments, including antigenic targets for *Shigella* vaccines and AMR data for revised treatment guidelines.

## Methods

### Study design and participants

We conducted a longitudinal cohort study with a 9·5-month follow-up at the Chainda South Health Facility in Lusaka, Zambia, which provides outpatient care services to the residents of Kalikiliki and Mtendere East (Fig. 1). These communities are home to over 25,000 residents. They comprise sprawling peri-urban settlements with unplanned housing, poor WASH conditions, and high unemployment rates. Children under five years were enrolled in the study if their guardians provided informed consent, lived within the catchment area, and were willing to visit the health facility for each diarrheal episode and provide diarrheal stool samples for testing.

**Figure 1.**
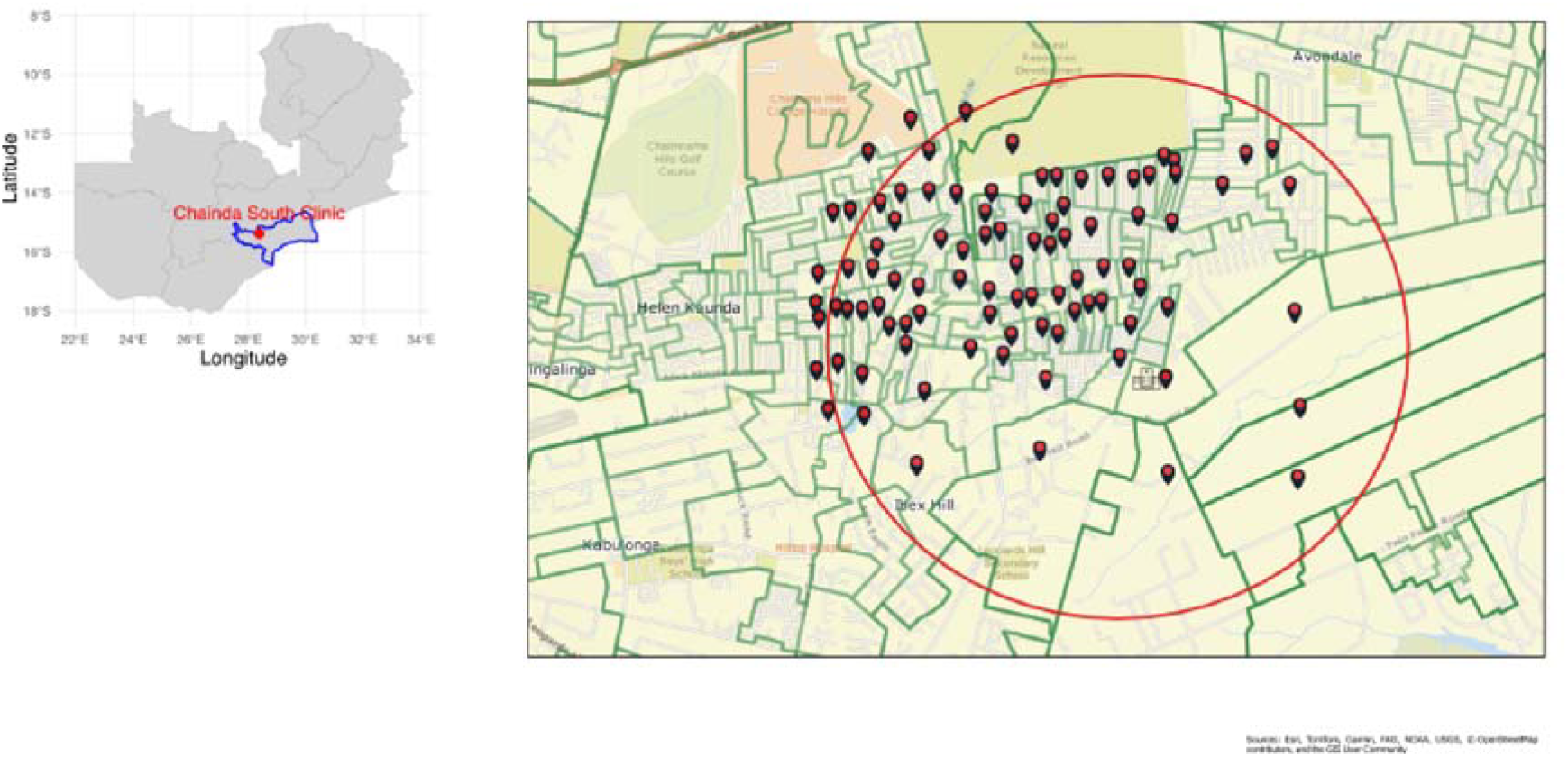
shows the map of Zambia with the administrative boundaries for Lusaka Province (in blue) and the location of Chainda South Clinic (latitude/longitude -28·35695/-15·3903) in red. On the right is the catchment area for Chainda South Clinic on MapsWithMe, showing the ‘Egg Yolk’ sampling method used during the census. The red pins represent standard census enumeration areas (SEAs) of 150 households. The green lines represent the boundaries of the standard census areas, and the red circle represents a radius of 5 km around the health facility.

**Figure 2.**
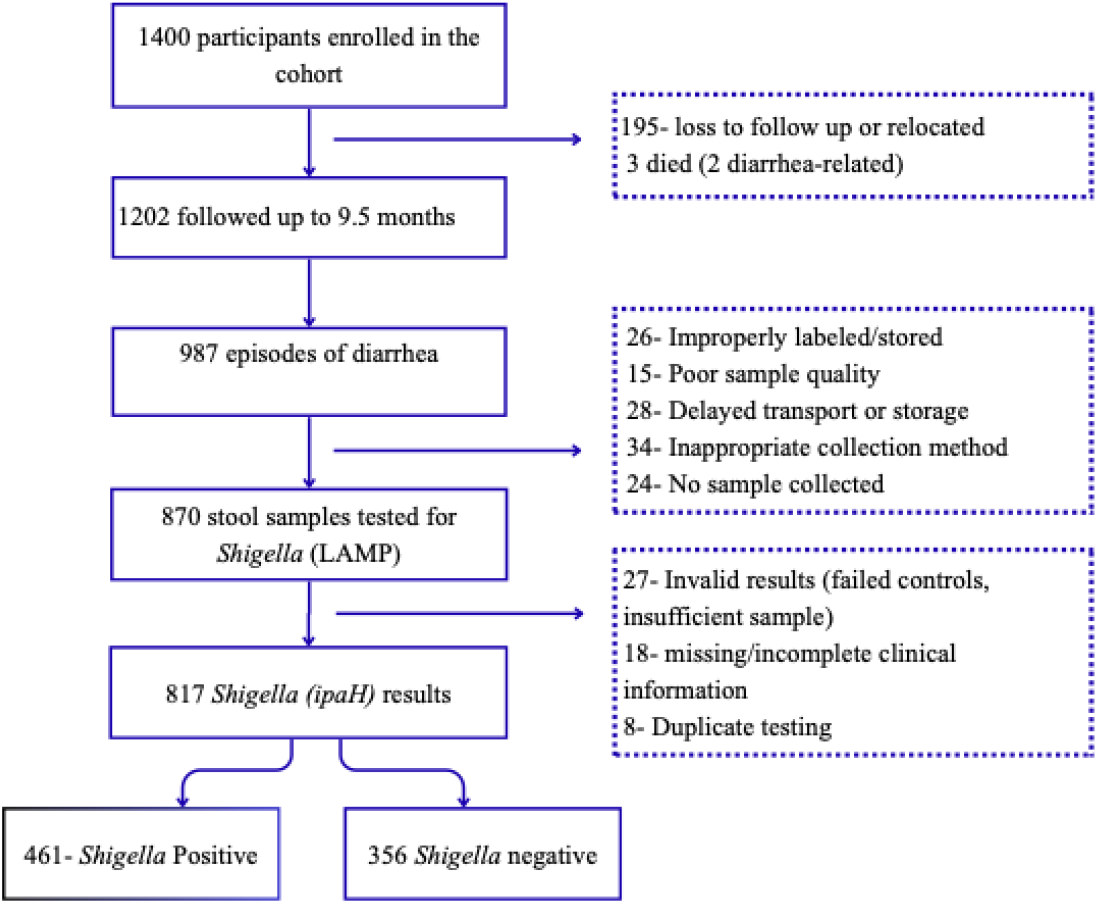
The study flowchart

### Ethical Considerations

The University of Zambia Biomedical Research Ethics Committee (UNZABREC) granted ethical approval (REF.NO. 696-2020), and the Zambian National Health Research Authority (NHRA) provided the final authorization for the study. Informed consent was obtained from all participants, and all data were anonymized.

### Study procedures

#### Enrollment

We conducted a community census to recruit and enroll participants by ‘egg-yolk/fried-egg’ sampling (i.e., selecting households within a 5 km radius of the study clinic (the central “yolk” area), to ensure that participants lived sufficiently close to likely visit the clinic for care. We used MapsWithMe (https://github.com/mapsme/api-ios) to help research assistants identify the geographic boundaries for the census (Fig. 1). Participants were enrolled if they were under five years old, lived in the catchment area of the study health facility, and their parents or guardians provided informed consent and agreed to present the child to the facility for any diarrhea. Children born after the census were not included.The primary outcome of the study was *Shigella*-positive diarrhea in children under five in the catchment areas of Chainda South Clinic, particularly in Kalikiliki and Mtendere East. Secondary outcomes included genomic characterization of *Shigella* isolates collected during the study, including species identification, genotypic AMR profiling to detect known AMR genes, and phenotypic antimicrobial susceptibility testing.

#### Diarrheal disease surveillance

A diarrhea episode was defined as having at least three loose stools within 24 hours or any loose stool with visible blood. A new episode was defined as an episode occurring after seven consecutive diarrhea-free days. A *Shigella-positive* case was defined as a diarrheal stool that tested positive for ipaH using the rapid loop-mediated isothermal amplification (LAMP) based diagnostic test (RLDT). After enrolment, passive surveillance for MSD was conducted at the health facility. Active surveillance was also carried out through monthly home visits to identify cases of diarrhea that might not have been reported to the clinic. Diarrheal severity was assessed using the Vesikari score and an in-house severity tool, the CIDRZ scoring system.^9^ All cases of gastroenteritis were managed according to national treatment guidelines, with ciprofloxacin recommended for treatment of *Shigella*-confirmed gastroenteritis (https://www.moh.gov.zm/?wpfb_dl=32).

#### Laboratory methods

##### a) Collection of stool samples for diarrheal cases

10-50 grams of stool were collected in the clinic in labeled, sterile stool collection bottles, stored in an ice-packed cooler at 2-8°C, and transferred to the lab within 2-4 hours. Stool was immediately cultured on arrival and then aliquoted and frozen at -80°C. For home collections, the guardian would collect stool and store it at cool temperatures for 1-2 hours until collection by a driver, where it was cultured, aliquoted, and stored at -80°C.

##### b) *Shigella* detection

Fecal samples were tested for *Shigella* (ipaH) using a rapid loop-mediated isothermal amplification (LAMP) based diagnostic tool (RLDT) targeting the ipaH gene, as described by Chakraborty et al.^10^ This semi-quantitative method incorporates bacterial load, with a threshold set at fluorescence >4000 RFU within 40 minutes time-to-result (TTR), preferentially selecting high-load infections and minimizing inclusion of low-level carriage. The samples were run alongside negative and positive controls.

##### c) Isolation, culture, and identification of *Shigella spp*

All diarrheal stool samples collected were enriched in Selenite-F broth and incubated at 37°C for 24 hours in an aerobic environment within four hours of collection, then subcultured onto MacConkey (Himedia, Mumbai, India), Xylose-Lysine Deoxycholate (XLD) (Himedia, Mumbai, India), and *Salmonella*-*Shigella* agar (SSA) (Himedia, Mumbai, India). Colonies suggestive of *Shigell*a (non-lactose fermenters) were subcultured onto nutrient agar. Identification was confirmed using biochemical tests, including the Lysine Iron Agar, Triple Sugar Iron Agar, SIM (Sulfur Indole Motility), and urea and citrate utilization tests. S*higella* presumptive isolates were confirmed using PCR targeting the ipaH gene to detect *Shigella*/EIEC. The isolates were serotyped as *S. flexneri, S. sonnei*, and *Shigella spp*. The primers are shown in Supplementary Table S1. *Shigella* isolates were stored in 15% glycerol at - 80°C for further testing.

##### d) Phenotypic antimicrobial susceptibility testing (AST)

Phenotypic antimicrobial susceptibility testing was conducted for *Shigella* isolates using the Phoenix™ Automated Microbiology System (BD Diagnostics, Sparks, MD, USA). Results were interpreted using the Clinical and Laboratory Standards Institute (CLSI M100) cut-off values. Multidrug-resistant (MDR) strains were defined as isolates exhibiting resistance to at least one drug from three or more antimicrobial classes.^11^

##### e) Genomic sequencing and analysis of *Shigella* isolates

Genomic DNA was extracted from revived isolates using the Quick-DNA^™^ Fungal/Bacterial Miniprep Kit (Zymo Research, Irvine, CA, USA). The extracted DNA was then shipped on dry ice to Latvia MGI Tech Laboratories for sequencing.

Whole-genome sequencing was performed on the extracted genomic DNA using the MGI DNBSEQ-G400 platform (1GB per sample). Adapters and low-quality reads (Phred score of < 30) were removed using Fastp.^12^ Read quality was determined using FastQC (https://www.bioinformatics.babraham.ac.uk/projects/fastqc/) and MultiQC.^13^ De novo assembly was performed using Shovill, with Spades as the assembler (https://github.com/tseemann/shovill?tab=readme-ov-file). Assembled reads were annotated in Prokka software^14^, and known AMR genes were screened using AMRfinderPlus (v. 3.11.26)^15^ using the *Escherichia* option to detect genes and point mutations. Virulence profiling was performed using the VirulenceFinder database on the Abricate tool (https://github.com/tseemann/abricate). We used MOB-suite^16^ to assemble plasmids and screened them for AMR genes as above.

All *Shigella* sequences were serotyped *in silico* using ShigaTyper v 2.0.5^17^, while lineage typing for *S. sonnei* was performed using Mykrobe (v0.13.0) (https://github.com/Mykrobe-tools/mykrobe?tab=readme-ov-file). The pan-genome was assessed in Panaroo^18^, and the core genome alignment was used to detect the single-nucleotide polymorphisms (SNPs) using SNP-sites v2.5.1.^18^ A maximum likelihood (ML) phylogeny for each species was built in IQ-TREE (v2)^19^ and visualized in the Interactive Tree of Life (iTOL).^20^ Publicly available genomes were incorporated into the tree to provide a global context for our isolates (Supp. Data S1).

#### Data management and data quality

Data were collected using a structured questionnaire on tablets programmed with Open Data Kit (ODK) software and configured on a secure server. After obtaining consent, research assistants conducted face-to-face interviews with the guardian in English or a local language (Nyanja/Bemba). Each participant was assigned a unique ID. The data collected included sociodemographic, WASH practices, healthcare-seeking behaviors, recent illnesses, nutritional status, and other potential risk factors for diarrheal disease. Child age, sex, and HIV-exposure/status were collected from the child’s Under-five health card. Clinical data were collected on electronic case report forms (CRFs) configured on the DHIS2 platform and included the type and duration of diarrhea, prior antibiotic use, and presenting symptoms. To ensure data quality, bench surfaces were disinfected between batches, sample IDs were tracked using barcodes, and PCR assays were run in duplicate with controls. About 10% of the data was double-entered to ensure transcription integrity.

### Outcomes

#### Sample size calculation and statistical analysis

We calculated a sample size of 1,334 participants for the surveillance cohort to generate a two-sided 95% confidence interval with a margin of error of 0·025. We assumed a hazard rate (λ) of 0·04 for confirmed *Shigella/EIEC* MSD cases based on MAL-ED data^21^, which reported *Shigella* incidence of ∼26 cases/100 child-years. Baseline characteristics were summarized as proportions, medians, and interquartile ranges (IQRs) for categorical and continuous variables. We used the Kaplan-Meier method to estimate the cumulative incidence of MSD with confirmed *Shigella/EIEC* etiology after a nine-and-a-half-month follow-up. We developed a parsimonious Cox proportional model by identifying factors independently associated with *Shigella*/EIEC incidence. Variables were limited to those with <5% missing data and were selected based on biological plausibility, prior literature evidence, and significance in the univariable analysis (p<0·05). When collinear, we retained the variable with stronger biological plausibility and literature support. Model performance and parsimony were evaluated using Akaike’s Information Criterion (AIC). Because most households contributed only one eligible child, we did not include a random effect for household; instead, we applied robust standard errors clustered at the household level as a conservative adjustment. Communities were designated as strata to account for differing baseline hazards. The proportional hazards assumption was assessed using Schoenfeld residuals. Potential bias was minimized by prespecifying eligibility criteria, conducting community sensitization to reduce differential participation, conducting interviews in a private space, using a standardized definition of diarrhea, employing SOPs for stool collection and laboratory testing, blinding laboratory staff to clinical data by maintaining separate databases for the clinic and laboratory. All analyses were performed using Stata 17 MP4 (StataCorp, College Station, TX, USA), and graphs were generated using the ggplot2 package in R software.

## RESULTS

A total of 1,400 children aged 0 to 5 years from 1,148 households were enrolled and followed between September 2020 and November 2021 (Table 1). 50·4% (n=705/1400) were female, and 50%(n=701/1400) were aged below two years. A high prevalence of stunting was observed at baseline (45·7%, n=640), and a small proportion (4·9%, n=66) of children were HIV positive. The average household size was 5·1 (SD = ±1·9). Most households had unimproved WASH facilities (88.7%, n=1018). The study reported three deaths, two of which were diarrhea-associated.

**Table 1.**
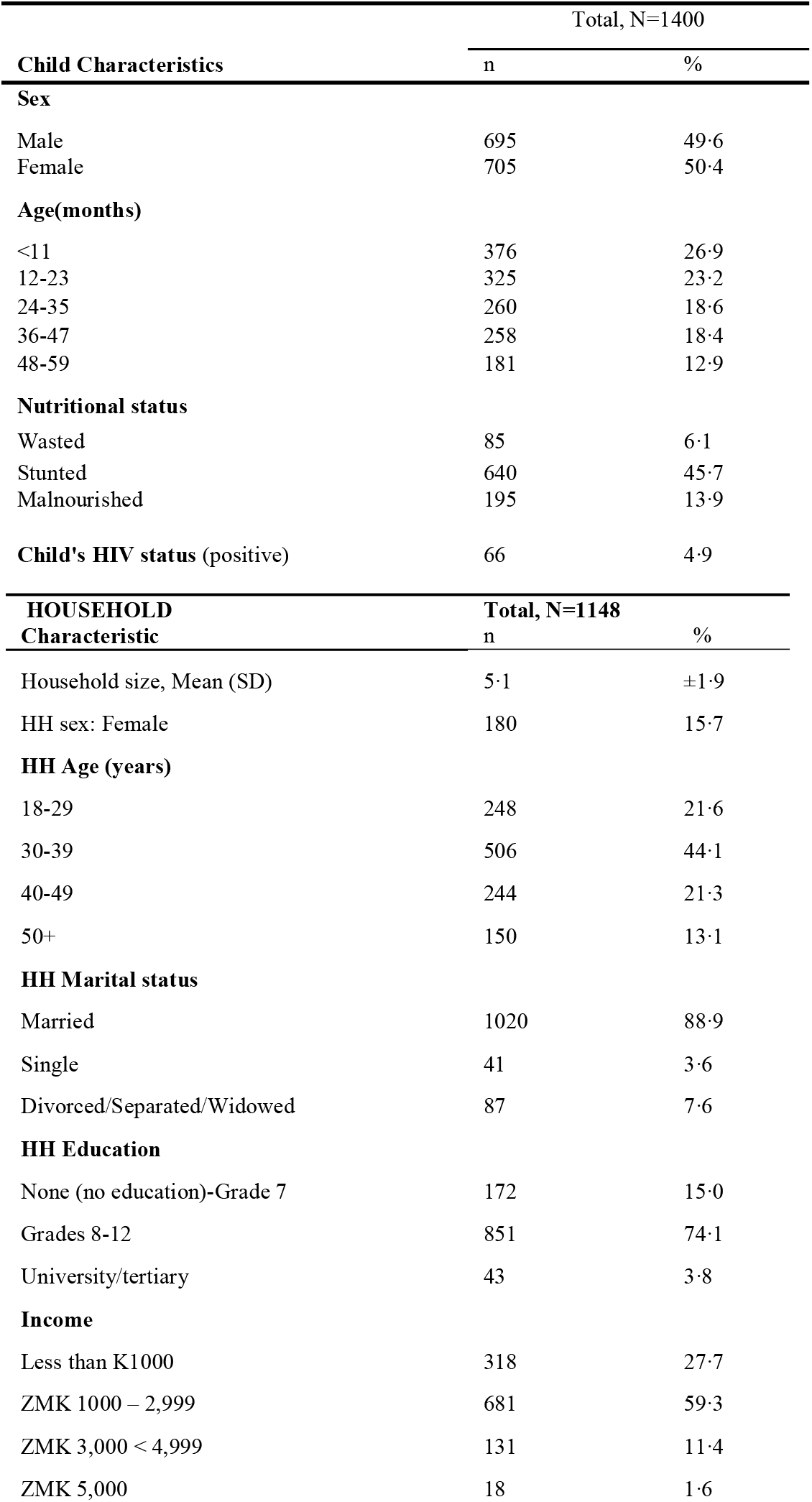
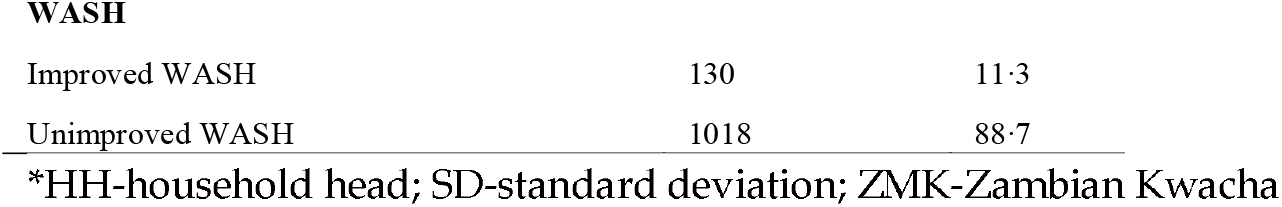
Baseline demographic characteristics of households and participants.

| Child Characteristics | Total, N=1400 |  |
| --- | --- | --- |
|  | n | % |
| **Sex** |  |  |
| Male | 695 | 49.6 |
| Female | 705 | 50.4 |
| **Age(months)** |  |  |
| <11 | 376 | 26.9 |
| 12-23 | 325 | 23.2 |
| 24-35 | 260 | 18.6 |
| 36-47 | 258 | 18.4 |
| 48-59 | 181 | 12.9 |
| **Nutritional status** |  |  |
| Wasted | 85 | 6.1 |
| Stunted | 640 | 45.7 |
| Malnourished | 195 | 13.9 |
| **Child's HIV status (positive)** | 66 | 4.9 |
| **HOUSEHOLD Characteristic** | **Total, N=1148** |  |
|  | n | % |
| Household size, Mean (SD) | 5.1 | $\pm 1.9$ |
| HH sex: Female | 180 | 15.7 |
| **HH Age (years)** |  |  |
| 18-29 | 248 | 21.6 |
| 30-39 | 506 | 44.1 |
| 40-49 | 244 | 21.3 |
| 50+ | 150 | 13.1 |
| **HH Marital status** |  |  |
| Married | 1020 | 88.9 |
| Single | 41 | 3.6 |
| Divorced/Separated/Widowed | 87 | 7.6 |
| **HH Education** |  |  |
| None (no education)-Grade 7 | 172 | 15.0 |
| Grades 8-12 | 851 | 74.1 |
| University/tertiary | 43 | 3.8 |
| **Income** |  |  |
| Less than K1000 | 318 | 27.7 |
| ZMK 1000 – 2,999 | 681 | 59.3 |
| ZMK 3,000 < 4,999 | 131 | 11.4 |
| ZMK 5,000 | 18 | 1.6 |
| WASH |  |  |
| Improved WASH | 130 | 11.3 |
| Unimproved WASH | 1018 | 88.7 |

\*HH-household head; SD-standard deviation; ZMK-Zambian Kwacha

### Clinical presentation of *Shigella*-positive diarrheal episodes

Over 9·5 months, 987 cases of MSD were identified at the health facility, and 870 diarrheal samples were tested for *Shigella*. 817 samples had valid, interpretable results for *Shigella* testing. Among the children who provided these samples, only 1·2% (n=10/817) presented with bloody diarrhea, while 87·2% (n=712/817) had watery diarrhea, and 6·0% (n=49/817) had mucoid diarrhea (Table 2). S*higella* (ipaH) was detected in 461/817 (56·4%) diarrheal stools using the LAMP method. It was detected in over half of watery diarrhea (56·6%, n=403/712), in 70% (n=7/10) of bloody diarrhea cases, and in 65·3% (n=32/49) of mucoid diarrhea cases. Diarrhea severity scoring indicated severe diarrhea in 3·2% (n=26/817) and 5·5% (n=45/817) using modified Vesikari and CIDRZ scores, respectively. Among the severe episodes, 15/26 (57·7%) and 30/45 (66·7%) tested positive for Shigella (ipaH) using the Vesikari and CIDRZ scores, respectively. 777/817 (95·1%) of diarrhoeal samples were available for culture. The culture method detected 83/777 (10·7%).

**Table 2.** Clinical characteristics of *Shigella-positive* (ipaH) diarrhea cases.

|  | Total<br>(N = 817) | *Shigella* Positive<br>(n = 461, 56.4%) |
| --- | --- | --- |
| Characteristic | n (% total) | n (row %) |
| **Type of Diarrhoea** |  |  |
| Watery | 712 (87.2) | 403 (56.6) |
| Bloody | 10 (1.2) | 7 (70.0) |
| Mucoid | 49 (6.0) | 32 (65.3) |
| Rice water | 1 (0.1) | 1 (100) |
| Loose stool | 45 (5.5) | 18 (40.0) |
| **Able to feed** |  |  |
| No | 111 (13.6) | 59 (53.9) |
| **History of fever** |  |  |
| Yes | 253 (31.0) | 142 (56.1) |
| **Hydration Status** |  |  |
| No dehydration | 779 (95.8) | 442 (56.7) |
| Some dehydration | 31 (3.8) | 17 (54.8) |
| Severe dehydration | 3 (0.3) | 2 (66.7) |
| Missing | 4 (0.5) | 0 |
| **Abdominal pains** |  |  |
| Yes | 294 (36.0) | 172 (58.5) |
| **Vomiting** |  |  |
| Yes | 125 (15.3) | 69 (55.2) |
| **Nutritional status** |  |  |
| wasted (WAZ < -2.0) | 73 (8.9) | 36 (49.3) |
| Stunted (HAZ < -2.0) | 407 (49.8) | 228 (56.1) |
| Malnourished (WHZ < -2.0) | 204 (25.0) | 100 (49.0) |
| **Severity scoring** |  |  |
| **Modified Vesikari** |  |  |
| Mild (<8) | 668 (81.7) | 371 (55.5) |
| Moderate (8–10) | 123 (15.1) | 75 (61.0) |
| Severe (≥11) | 26 (3.2) | 15 (57.7) |
| **CIDRZ score** |  |  |
| Mild/moderate (<5) | 772 (94.5) | 431 (55.8) |
| Severe (≥5) | 45 (5.5) | 30 (66.7) |

#### *Shigella* incidence and associated risk factors

We next described the incidence and potential risk factors for *Shigella*-positive diarrhea (Table 3). During the period under review, participants collectively contributed 9581·7 child-months of follow-up time. The overall incidence of *Shigella* was 24·0 (95% CI 21·1-27·3) per thousand child-months. The highest incidence was observed in children aged 2-3 years, with rates of 30·5 (95% CI: 24·0-38·6) and 29·6 (95% CI: 22·5-38·8) for two and three-year-olds, respectively. Children aged 2-3 years had a significantly higher risk of infection than those under one year, and the risk was significantly reduced in the 4–5-year-olds, and the log-rank test indicated a significant difference in cumulative incidence among age groups, χ^2^(3) = 15·65, p = 0·0013 (supplementary Fig. 5). Improved WASH had significantly reduced the risk of infection compared to non-improved WASH (AHR 0·39: 95% CI (0·19-0·77). Households with electricity showed a reduced risk of infection in the univariable but not the multivariable analysis (AHR 0·77: 95% CI (0·57-1·04)). Malnutrition was likewise significantly associated with *Shigella* infection in the univariate but not the multivariate analysis (AHR 1·30: 95% CI (0·91-1·86).

**Table 3.**
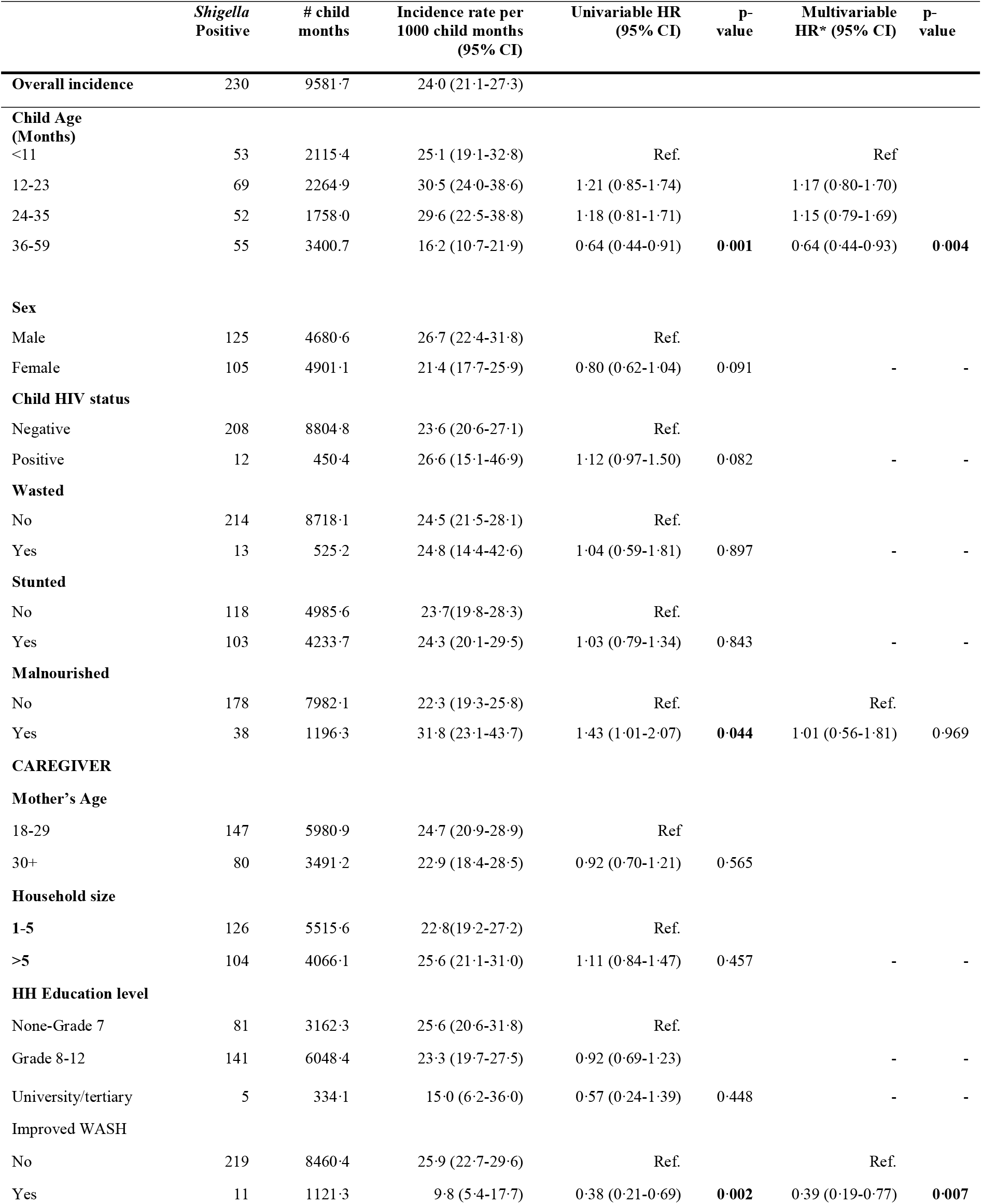
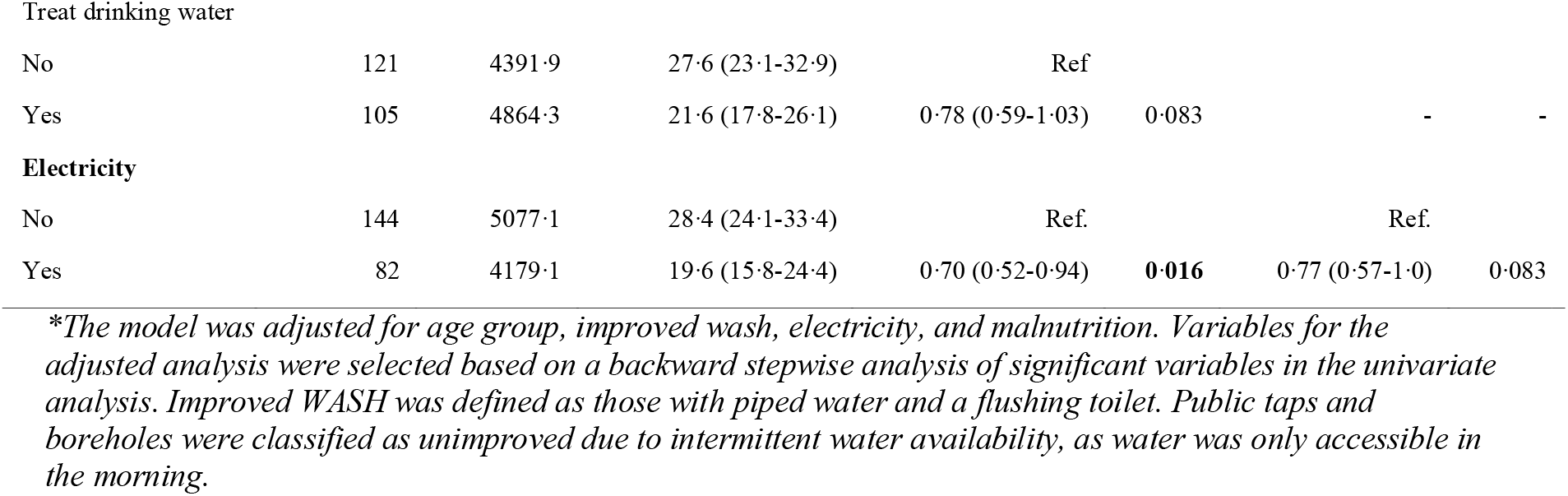
Incidence and Risk Factors of *Shigella* Infection.

#### Genomic characterization of the *Shigella*

Next, we were interested in the genomic makeup of locally circulating *Shigella* strains. Out of 83 culture-positive isolates, 75 isolates were revived for WGS. Of the 75, 69·3% (n=52/75) *Shigella* isolates met the quality threshold and were evaluated using WGS. Using Shigatyper, we identified 22 (42·3%) isolates belonging to three *Shigella* serotypes: 10 *S. flexneri*, 9 *S. sonnei*, 3 *S. boydii*, and none of *S. dysenterie* (Fig.3 and Fig.4). *19* isolates (36·5%) were identified as Enteroinvasive *E. coli* (EIEC), while 11 (21·2%) were classified as not *Shigella* or EIEC, possibly due to the quality or completeness of the sequence data. Phylogenetic analysis of *S. flexneri* revealed the study isolates belonged to three previously described phylogroups^22^, three in phylogroup 1 (PG1), five isolates in PG3, and two isolates in PG2 (Fig. 4). Phylogenetic analysis of *S. sonnei* indicated that all isolates belong to the widespread Lineage III (Fig. 4), described in previous research from Europe.^23^ The analysis revealed two *S. sonnei* clusters with distinct plasmid and virulence profiles. Shigatyper identified these clusters as the virulent *S. sonnei* form I(+) and the avirulent *S. sonnei* form II(-). The *S. boydii* analysis revealed that all three *S. boydii* isolates belonged to the previously described phylogenetic clade II (Fig. 3).^24^

**Figure 3.**
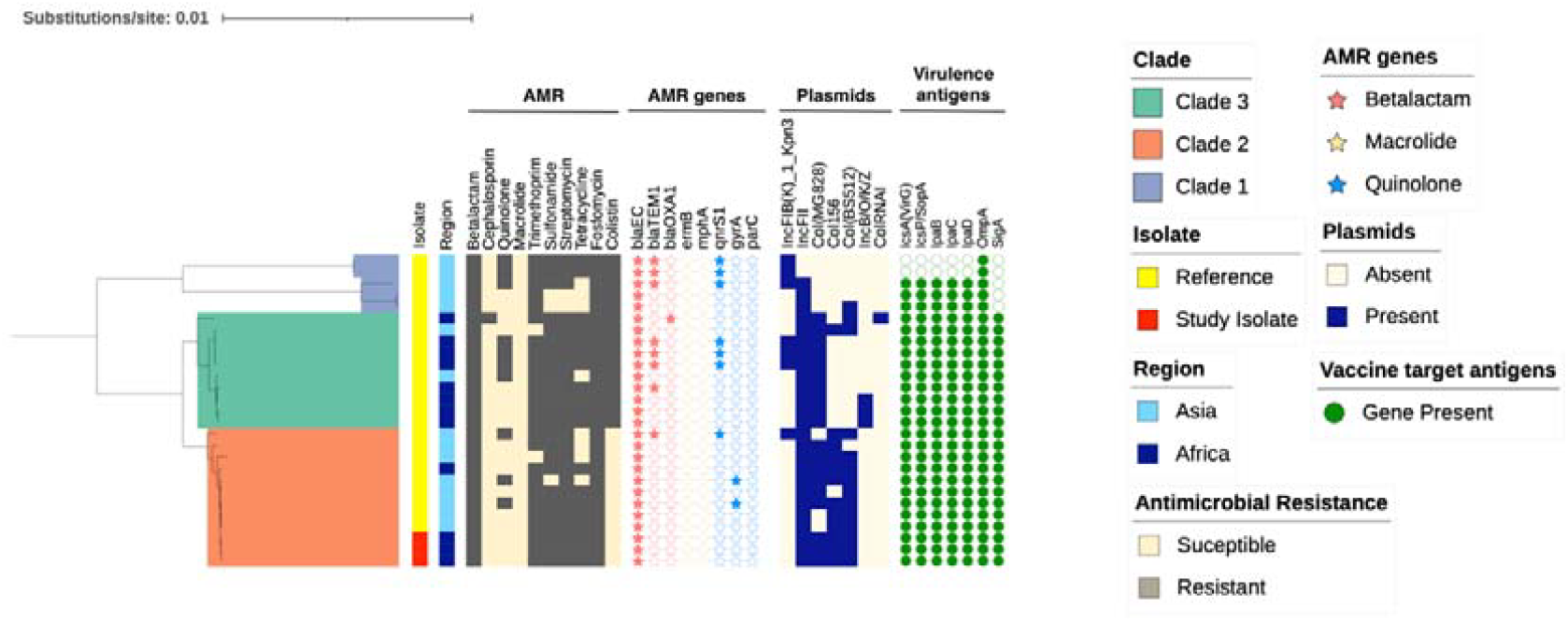
represents the phylogenetic tree showing the genomic characteristics of S. boydii isolates. Included are 3 study isolates alongside 24 publicly available reference genomes. The tree was midpoint-rooted, and study isolates were identified to belong to clade 2. The genomic metadata details are outlined in the legend and include antibiotic class (with gray indicating resistance), the presence of AMR genes for beta-lactams (red), macrolides (yellow), and quinolones (blue), the presence of plasmids (blue) and vaccine target antigens (green circle).

**Figure 4.**
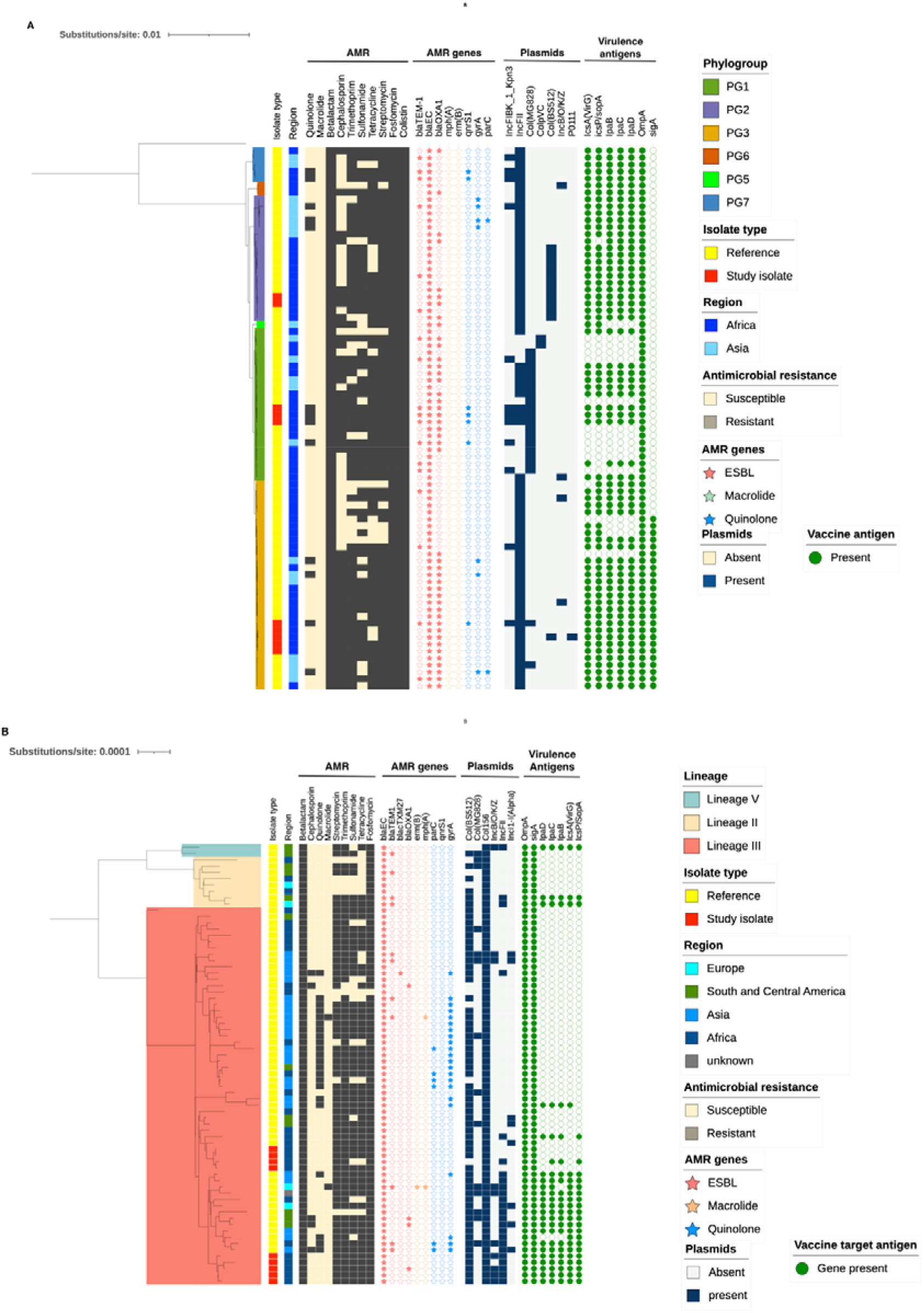
Phylogenetic trees for *S. flexneri (A)* and *S. sonnei (B)* isolates. A) Depicts the phylogeny of S. flexneri, including 10 study isolates and 70 publicly available reference genomes, rooted using an E. coli genome. The phylogeny suggests the study isolates belong to three *S. flexneri* phylogroups: PG1 (serotype 1b, n=3), PG2 (3a, n=2), PG3 (2a, n=5). B) Represents the mid-point rooted phylogeny of *S. sonnei* isolates, including 9 study isolates and 61 reference genomes. The outer columns on both trees display genotypic and metadata information, showing AMR-resistant (gray) and susceptible (cream) colors; plasmid presence (dark blue) and absence (light blue); and the presence of virulence genes (filled green circles) and absence (unfilled circles).

### Phenotypic and genomic antimicrobial resistance of *Shigella* isolates

To gain insight into the susceptibility of *Shigella* isolates to first- and second-line antibiotics, we tested the phenotypic AMR for relevant antibiotic classes among the 22 isolates identified as *Shigella* from WGS. The most common resistance was against trimethoprim-sulfamethoxazole (77·3%, n=17), ampicillin (72·7%, n=16), cefazolin (63·6%, n=14), and gentamicin (40·9%, n=9). Ertapenem resistance was observed in only one *S. flexneri*. Resistance to ciprofloxacin was observed solely in *S. flexneri* isolates (40%, n=4), and no resistance to azithromycin was observed. Multi-drug resistance was identified in 63·6% (n=14) of the isolates, with the highest proportion observed in *S. flexneri* (80%, n=8) (see Table 4).

**Table 4.**
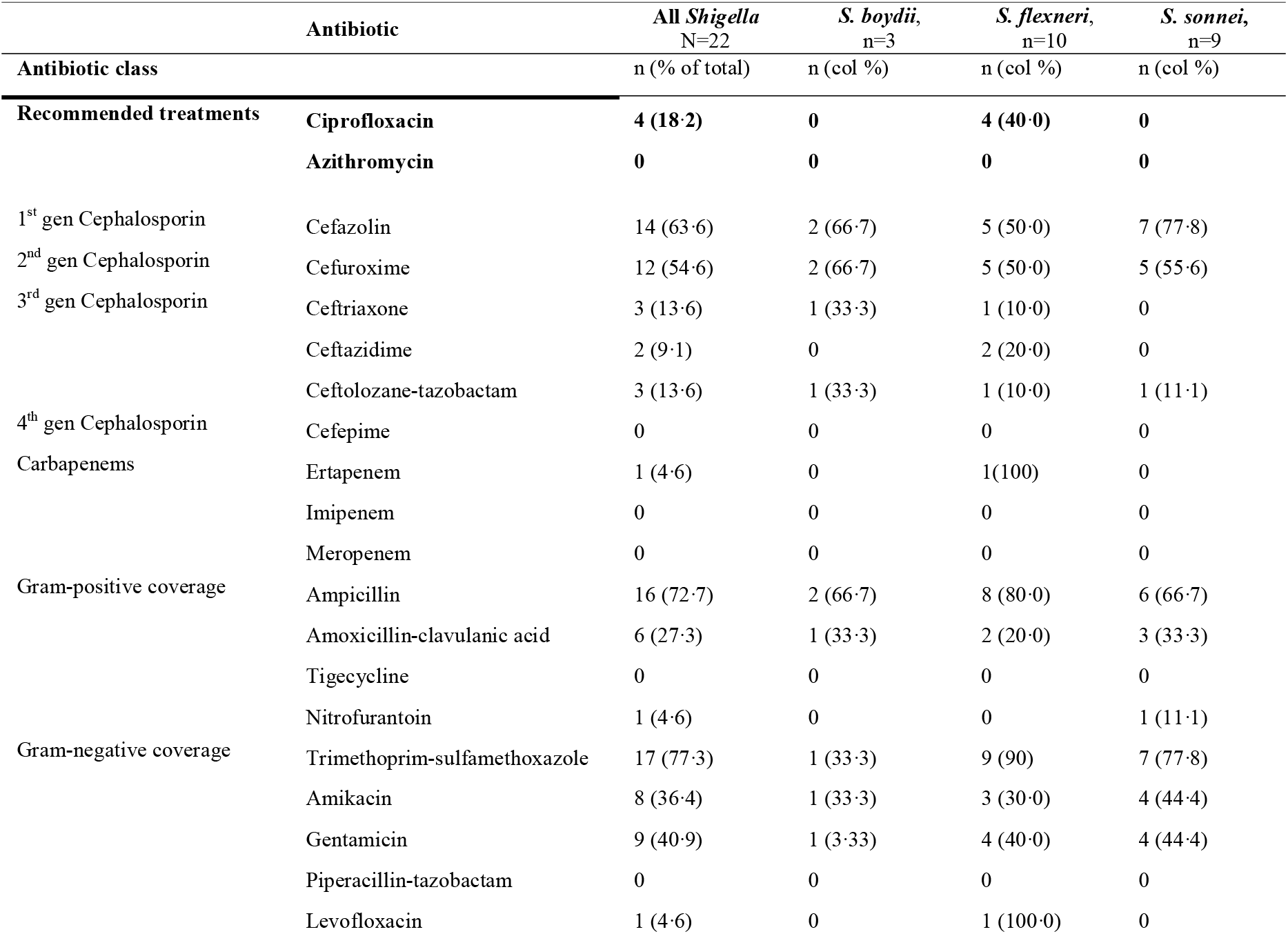
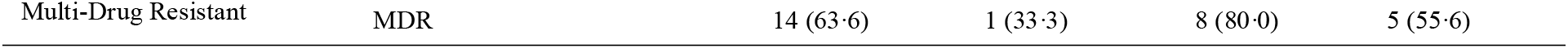
Phenotypic antimicrobial resistance profiles for *Shigella* isolates.

To understand the genomic basis of antimicrobial resistance (AMR) in *Shigella* isolates, we identified the presence of known AMR determinants. Resistance genes for trimethoprim, sulfonamides, tetracyclines, aminoglycosides, and beta-lactams were observed among all *Shigella* isolates, while resistance genes for other antimicrobial classes varied by serotype (Fig. 5). The key AMR genes identified included *dfrA1* (trimethoprim), *blaEC* (beta-lactam), *sul1* (sulfonamide), and *aac(6’)-Ib* and *aadA1* (aminoglycoside). The *S. flexneri* isolates had the largest number of AMR genes.

**Figure 5.**
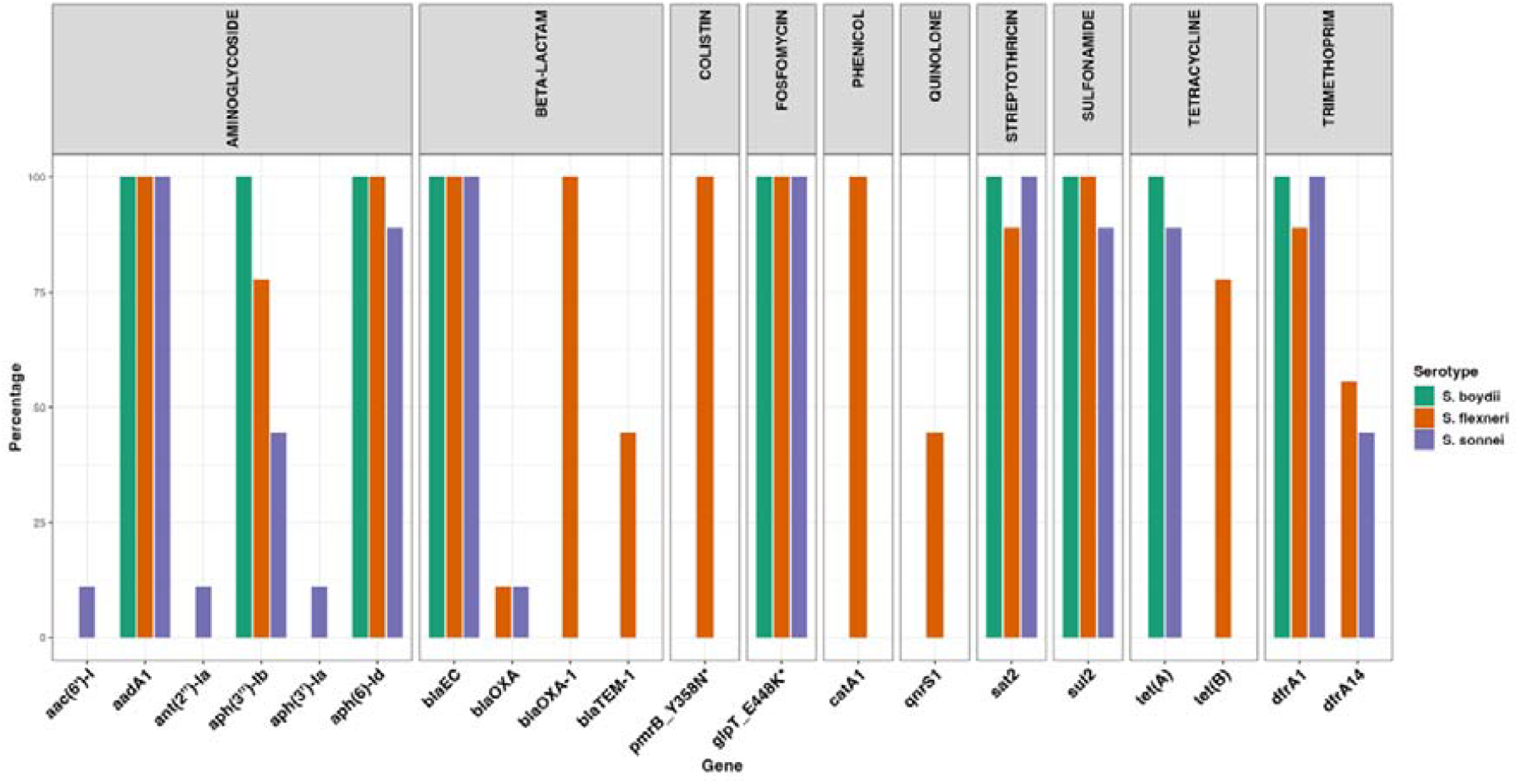
shows the AMR profiles and the distribution of AMR genes in the study isolates. The bars are grouped by AMR class (displayed on top) with genes indicated on the x-axis and colored according to the *Shigella* serotype. The bar plots represent the percentage (%) of AMR determinants detected for S. flexneri (10 isolates), *S. sonnei* (9 isolates), and *S. boydii* (3 isolates). Genes with an asterisk “*” indicate resistance conferred by point mutations.

We then examined resistance to macrolides and quinolones, the recommended first-line treatments for Shigellosis. We observed that 40% (n=4) of *S. flexneri* isolates carried the *qnrS1* gene, which typically encodes quinolone resistance on plasmids or mobile genetic elements (MGEs). The detection of *qnrS1* corresponded with phenotypic resistance to ciprofloxacin in 75% (n=3/4) isolates. No chromosomal mutations in the quinolone resistance-determining regions (QRDR) of the gyrA and parC genes or genes encoding macrolide resistance were detected in the study isolates. Beta-lactamase genes were observed in multiple isolates, with *blaOXA-1* and *blaTEM-1* identified only in *S. flexneri* isolates (*blaOXA-1*-100%, *blaTEM-1*-40%) (Fig. 5). The detection of these beta-lactamase genes corresponded to phenotypic resistance to cephalosporins but not carbapenems. *blaCTX*-associated resistance, including the *blaCTX-M-27* and *blaCTX-M-15* genes reported in *Shigell*a outbreaks in Europe ^25^, was not detected in this study.

Further analysis of the concordance between genotypic and phenotypic AMR profiles revealed a largely concordant relationship. However, five isolates showed no phenotypic resistance to trimethoprim-sulfamethoxazole, despite the presence of the *sul2* and *dfrA* genes, while one isolate demonstrated phenotypic resistance but lacked the corresponding gene for ertapenem. Additionally, 12 isolates exhibited no resistance to amikacin and gentamicin, even with *aadA1, aph(3’)-Ia*, and *aph(6)-Shigella* AMR determinants and virulence factors are known to be carried on plasmids, facilitating horizontal gene transfer between bacteria.^26^ Therefore, we screened the study isolates for known plasmids. Our analysis shows that all *S. sonnei* and *S. boydii* presented with Col156 and 89% and 100% with Col(BS5120), respectively. In contrast, all *S. flexneri* and *S. boydii* isolates presented with IncFII_1, while 40% of *S. flexneri* presented with IncFIB(K) plasmids (Fig. 3, 4). Further analysis showed that the *qnrS1* gene, associated with reduced quinolone susceptibility, was carried exclusively on the IncFIB(K) plasmid, which harbored multiple other AMR genes (supplementary Fig. 3). The large plasmid, IncB/O/K/Z, known to carry an array of ESBL genes^27^, was identified in 4/9 *S. sonnei* isolates but carried no ESBL genes. Other plasmids were observed less frequently.

We also evaluated whether the *Shigell*a isolates carried and likely expressed seven O-specific lipopolysaccharide (OSP) genes used in candidate vaccines in phases I and II of clinical trials.^28^ Our findings indicate that all seven antigens were present in all isolates except for sigA, which was absent in *S. flexneri’s* PG1 and PG3. Additionally, virulence plasmid genes (virG, icsP, ipaB, ipaC, and ipaD) were absent in one *S. sonnei* cluster (Figs. 3 and 4).

## Discussion

We report a high incidence of moderate to severe diarrheal disease with *Shigella* in children under five (24·1 per 1000 child months), with the highest incidence in children aged 2-3 years. This incidence is higher than that reported in the GEMS study conducted at African and Asian sites^4^ but comparable to results from the quantitative molecular re-analysis of the MAL-ED study.^21^ The key risk factors for *Shigella/EIEC* infection identified included age and source of drinking water, with higher risks observed in children 2-3 years old and participants from households that didn’t have water piped into the house or yard. Being underweight and household electricity were significant risk factors only in the unadjusted model, likely reflecting confounding or collinearity with other socioeconomic factors, such as unimproved WASH, that may influence both pathogen exposure and malnutrition. The findings align with previously reported risk factors for *Shigella/EIEC* in a pooled analysis from 23 LMICs.^29^ Our study detected *Shigella/EIEC* in 56·6% of watery stool samples, indicating that *Shigella/EIEC* frequently presents as watery diarrhea. Current diarrhea treatment guidelines do not support empiric antibiotic treatment for *Shigella/EIEC* in children presenting with watery diarrhea. However, the Antibiotics for Children with Severe Diarrhea (ABCD) trial demonstrated that watery diarrhea with confirmed or presumed bacterial etiology resulted in shorter diarrhea duration and less hospitalization/death, as well as less growth faltering in children with *Shigella*, when treated with azithromycin.^30^ We detected *Shigella/EIEC* in 56·4% (461/817) vs 11·2% (83/741) of stool samples using PCR and culture methods, respectively. These results are consistent with a similar study from Burkina Faso, which reported 23·2% detection of *Shigella/EIEC* using culture vs 44·8% with similar LAMP equipment.^31^ PCR-based studies consistently report higher case detection rates than traditional culture methods.^21,32^ *Shigella* is notoriously difficult to culture due to its low infectious dose, rapid loss of viability in the environment (e.g, during transportation), fastidious growth requirements, and prior antibiotic treatment administered before presentation to the health facility may reduce culture positivity.^33^

Our WGS analysis revealed numerous (n=19/52, 36·5%) bacterial isolates to be EIEC and not Shigella. This suggests that our biochemical profiling did not sufficiently distinguish between *Shigella* and EIEC. It also highlights a limitation of targeting the ipaH gene, which is shared by both *Shigella* and EIEC. This was expected as biochemical profiles of *Shigella* and EIEC are similar, and isolate typing was based on PCR targeting the shared ipaH gene.^34^ Additionally, we only speciated *S. flexneri* and *S. sonnei*, while other *Shigella* serotypes were reported as *Shigella spp*. Our WGS detection of EIEC may explain the higher incidence reported in our study compared to the GEMS study, which used a TaqMan Array Card to detect 32 pathogens and differentiated true pathogen-attributable cases from incidental detection.^32^ Our genomic data, however, mirror the molecular subtyping findings from the MAL-ED study, which reported only 49% of ipaH-positive *Shigella* infections as *S. flexneri* and *S. sonnei*.^5^ This suggests that many ipaH-positive *Shigella* may be EIEC, since *S. flexneri* and *S. sonnei* account for most *Shigella* infections. To assess the potential influence of EIEC on disease incidence, we downweighted the ipaH-positive incidence by the WGS-derived EIEC proportion (36·5%), resulting in a WGS-adjusted *Shigella* incidence of 15·2 (11·5-18·9), declining from 24·0 (21·1-27·3) per 1000 child-months. More sensitive and affordable point-of-care molecular diagnostic tools are needed to detect *Shigella*, particularly in regions of high EIEC burden. Our results also highlight the importance of carefully considering the choice of diagnostic tests when reporting *Shigella* incidence and estimating the future value of interventions such as vaccines.

The WHO issued guidelines recommending quinolones and macrolides for treating *Shigella* in 2005. Since then, increased antibiotic resistance of up to 44% for ciprofloxacin has been reported in this pathogen in Southeast Asia.^35^ The AMR profiling of our study isolates identified multi-drug-resistant strains, including four out of ten *S. flexneri* isolates resistant to quinolones (ciprofloxacin). These findings align with the WHO’s classification of fluoroquinolone-resistant *Shigella* as a priority pathogen, emphasizing the need to continually re-evaluate national dysentery treatment guidelines and accelerate *Shigella* vaccine development as strategies to reduce antimicrobial use and address possible regional rises in quinolone resistance in *Shigella* spp. Nonetheless, macrolides remain an effective treatment option for *Shigella* in Zambia. The detection of the quinolone-resistant qnrS1 gene in *S. flexneri* isolates is consistent with findings of plasmid-mediated quinolone resistance (PMQR) reported in two of six *Shigella* isolates from neighboring Malawi, suggesting potential regional transmission. In both countries, the contigs harboring the *qnrS1* gene also contained the *blaTEM-1B, sul2, aph(3′′)-Ib, and aph(6)-Id* genes^36^, suggesting that these genes may be carried on an MDR plasmid. However, these results contrast with the *Shigella* WGS results from the GEMS and Vaccine Impact on Diarrhea in Africa (VIDA) studies, which document minimal quinolone resistance at the African sites.^3,37^ The discrepancy likely reflects temporal trends of rising quinolone resistance in sub-Saharan Africa, as earlier analyses were conducted on samples collected between 2007-2011 and 2015-2018, respectively. We observed no macrolide resistance, suggesting that azithromycin remains an effective therapy for *Shigella* diarrhea in Zambia. These findings show the need for ongoing genomic AMR surveillance to inform *Shigella* treatment strategies.

Plasmids play a major role in the virulence and AMR profiles of *Shigella*.^38^ They are also mobile and able to spread through horizontal gene transfer, contributing to the population-level spread of AMR.^39^ Our analysis identified mainly Inc-type and col-like plasmids. Inc plasmids are known to carry transfer/mobilization features like tra loci/oriT/relaxase, highlighting their capacity for horizontal gene transfer. The ciprofloxacin resistance-conferring qnrS1 gene was carried exclusively on the IncFIB(K) plasmid, which also carried multiple AMR genes. This plasmid has been associated with multidrug resistance (MDR) in *Shigella* in Malawi^36^, Tanzania^40^, and other Enterobacteriaceae, including *Klebsiella* and *Salmonella* in South Africa.^41^ Our findings underscore the potential for plasmids to contribute to AMR spread through horizontal transmission between species within SSA. However, unlike the previous study, no *blaCTX-M-*1 genes were detected on this plasmid. The coexistence of multiple AMR genes on a plasmid can accelerate the population-level spread of MDR, conferring simultaneous resistance to multiple antibiotic classes, which limits treatment options and increases healthcare burden costs.

The phenotypic and genotypic AMR profiling indicated some level of concordance. However, several discrepancies were identified between the phenotypic and genotypic AMR assessment profiles, highlighting the limitations of relying solely on genotypic profiling for AMR. These discrepancies are common. At the genomic level, they could be attributed to some AMR genes being non-functional, poorly expressed, or requiring specific promoters or mutations to confer resistance. At the population level, resistance genes may be carried by minority subpopulations or lost through plasmid instability during subculturing or storage. Phenotypic testing conditions or detection thresholds may also fail to capture low-level expression of resistance.^42^ Additionally, this analysis classified intermediate phenotypes as susceptible, potentially underestimating genotypic predictions. Nonetheless, our results emphasize the importance and benefits of monitoring AMR trends using integrated phenotypic and genotypic methods.

As *Shigella* becomes increasingly resistant to antimicrobials, developing and licensing safe, immunogenic, and efficacious vaccines against *Shigella* will become increasingly important. Several vaccines have demonstrated protective potential and are in advanced stages of development, including InvaplexAR-Detox, whose Phase Ia/b clinical trial is ongoing in Zambia; the S4V-EPA by LimmaTech; and AltSonflex1-2-3, a GMMA-based vaccine candidate by GlaxoSmithKline (GSK).^28^ Our analysis identified three *Shigella* serotypes with varying virulence antigens, including the absence of SigA in some *S. flexneri* isolates. This can be explained by known gene plasticity within *Shigella* serotypes, as genes are carried on MGEs, inactivated, or lost during selective pressure.^43^ This could impact vaccine coverage and highlights the need for a multivalent *Shigella* vaccine, targeting more conserved antigens to offer broad protection against *Shigella*, with or without AMR. To effectively mitigate *Shigella* infections, vaccine strategies can complement WASH improvements, as evidence from randomized trials indicates that household-level WASH interventions alone are insufficient. ^44,45^

### Strengths and limitations

The study was community-based, with active surveillance to improve case detection. The combination of phenotypic and genotypic AMR profiling provides a comprehensive picture of *Shigella* epidemiology in Lusaka, with direct implications for local treatment guidelines.

A key limitation of nucleic acid amplification tests including PCR and LAMP that target the ipaH gene is that ipaH is present in both *Shigella* spp. and EIEC. In EIEC-endemic settings like Zambia, this likely overestimates the true burden of *Shigella* by inflating *Shigella*’s attributable fraction. WGS and molecular platforms like the customized TaqMan Array card used in the Enterics for Global Health (EFGH) study^46^ which targets multiple *Shigella*-specific markers for *S. flexneri, S. sonnei* speciation, and co-pathogens, could improve the accuracy of burden estimates and provide a comprehensive view of *Shigella* incidence and AMR to guide vaccine planning in SSA and Asia. To minimize over-attributing disease to *Shigella*, the LAMP used a validated fluorescence cutoff (>4000 RFU) within 40 minutes to exclude likely low-level carriage. This approach parallels the Ct-based thresholds used in GEMS^32^ and MAL-ED^21^ to focus on clinically relevant infections with higher bacterial loads. Data on antibiotic use were collected at each episode. However, during the COVID-19 pandemic, healthcare-seeking behaviour declined, and as such, children could have been treated with informally obtained antibiotics for illnesses, potentially influencing our *Shigella* culture yield. Additionally, the incidence findings are reflective of a peri-urban population, with limited generalisability to other rural settings in Zambia or the wider region. We did not collect data on ethnicity or tribe, which may limit our ability to determine whether the findings differ across ethnic groups and could further affect the generalizability of the results to other populations. The sample size was calculated for incidence estimation without explicit adjustment for clustering or attrition. Follow-up was good, and clustering was minimal since a few households had more than one enrolled child. However, the study was not powered for secondary analyses, limiting the interpretation of risk factors with small effect sizes or subgroup comparisons. Not all isolates were successfully revived, limiting the representation of genomic data. The small study population and single study site also limit how well circulating sero and genotypes represent the broader population for vaccine development. Despite this, the study documents the genetic diversity of locally circulating *Shigella* species and serotypes.

## Conclusion

Our study reveals a significant burden of *Shigella* diarrhea in children under five in Lusaka, Zambia. It highlights the rising genotypic and phenotypic resistance of *Shigella* to quinolones in sub-Saharan Africa, emphasizing the urgent need for effective vaccines and strategies to reduce morbidity and mortality from the disease.

## Data Availability

All raw sequence reads from the study have been deposited in the ENA database at EMBL-EBI under project accession number PRJEB100814. The study data, including the metadata and accession numbers, have been deposited on figshare (10.6084/m9.figshare.28937558).

## Code availability

This study did not use any custom code. The methods section outlines all bioinformatic packages used.

## Author contributions

Conceptualization-RC, SB, CS, MS, VH, DM, and MC; Data verification-SS, CCL, FL, and MC; Access to raw data-MC, MS, SB, and GP; Data curation, formal data analysis, and Bioinformatics analysis-MC, SB, DM, VH, JB, and GP; Laboratory investigations-KM, KC, SS, CCL, and FL. Writing the original draft-MC and VH; editing and reviewing the final manuscript-CS, DM, MS, CCC, RC, SB, MS, VH, KM, KC, SS, CCL, and FL. All authors contributed to the writing and editing of the manuscript. All authors agreed to submit for publication.

Conflict of Interest: V.H. reports institutional funding support for this work by the Netherlands Organization for Health Research and Development (ZonMw) (VENI 09150161810022), Health-Holland/Top Sector Life Sciences & Health (GLORIA LSHM21033 and Track-AMR LSHM23007), the National Institutes of Health (NIH) (R01 AI173360), and the Wellcome Trust (219775/Z/19/Z); all payments went to the host institution. All other authors have no conflict of interest to disclose.

## Acknowledgments

We also sincerely thank the children, their guardians, and the neighbourhood health committees for their generosity and willingness to participate in this study. We thank the study staff, including the research clinicians and nurses, research assistants, laboratory staff, and community health workers at the Chainda South clinic, who made this study possible. We also acknowledge the support from the Enteric Diseases and Vaccines Research Unit (EDVRU) and Analysis (AU) Unit at the Center for Infectious Disease Research in Zambia (CIDRZ). We would also like to thank the team at the Center for Infectious Disease Control in the Netherlands for their support with the genomic analysis.

This work is part of the EDCTP2 program supported by the European Union (grant number RIA2018V-2308-ShigaPlexIM). It was also supported by The Schlumberger Foundation’s flagship program, The Faculty for the Future Foundation (FFTF), through the postgraduate fellowship awarded to Mwelwa Chibuye. We acknowledge funding from the Netherlands Organization for Health Research and Development (ZonMw) VENI 09150161810022 (V.C.H.), Health-Holland AMR-Global, Gloria, and Track-AMR (V.C.H. and D.R.M.). The funders had no role in the data collection, analysis, manuscript preparation, or decision-making process regarding publication.

